# Determinants of ascending aortic morphology: Cross-sectional deep learning-based analysis on 25,073 non-contrast-enhanced MRI of NAKO

**DOI:** 10.1101/2024.07.12.24310356

**Authors:** Louisa Fay, Tobias Hepp, Moritz T. Winkelmann, Annette Peters, Margit Heier, Thoralf Niendorf, Tobias Pischon, Beate Endemann, Jeanette Schulz-Menger, Lilian Krist, Matthias B. Schulze, Rafael Mikolajczyk, Andreas Wienke, Nadia Obi, Bernard C. Silenou, Berit Lange, Hans-Ulrich Kauczor, Wolfgang Lieb, Hansjörg Baurecht, Michael Leitzmann, Kira Trares, Hermann Brenner, Karin B. Michels, Stefanie Jaskulski, Henry Völzke, Konstantin Nikolaou, Christopher L. Schlett, Fabian Bamberg, Mario Lescan, Bin Yang, Thomas Küstner, Sergios Gatidis

## Abstract

**Background:** Pathologies of the thoracic aorta are associated with chronic cardiovascular disease and can be of life-threatening nature. Understanding determinants of thoracic aortic morphology is crucial for precise diagnostics and preventive and therapeutic approaches. This study aimed to automatically characterize ascending aortic morphology based on 3D non-contrast-enhanced magnetic resonance angiography (NE-MRA) data from the large epidemiological cross-sectional German National Cohort (NAKO) and to investigate possible determinants of mid-ascending aortic diameter (mid-AAoD).

**Methods:** Deep learning was used to automatically segment the thoracic aorta and extract ascending aortic length, volume, and diameter from 25,073 NE-MRAs. Descriptive statistics, correlation analyses, and multivariable regression were used to investigate statistical relationships between mid-AAoD and demographic factors, hypertension, diabetes, alcohol, and tobacco consumption. Additionally, automated causal discovery analysis using the Peter-Clark algorithm was performed to identify possible causal interactions.

**Results:** Males exhibited significantly larger mid-AAoD than females (M: 35.5±4.8 mm, F: 33.3±4.5 mm). Age and body surface area (BSA) were positively correlated with mid-AAoD. Hypertensive and diabetic subjects showed higher mid-AAoD. Hypertension was linked to higher mid-AAoD regardless of age and BSA, while diabetes and mid-AAoD were uncorrelated across age-stratified subgroups. Daily alcohol consumption and smoking history exceeding 16.5 pack-years exhibited highest mid-AAoD. Causal analysis revealed that age, BSA, hypertension, and alcohol consumption are possibly causally related to mid-AAoD, while diabetes and smoking are likely spuriously correlated.

**Conclusions:** Mid-AAoD varies significantly within the unique large-scale NAKO population depending on demographic factors, individual health, and lifestyle. This work provides a proof-of-concept for automated causal analysis which can help disentangle observed correlations and identify potential causal determinants of ascending aortic morphology.

**CLINICAL PERSPECTIVE:** Non-contrast-enhanced magnetic resonance angiography (NE-MRA) is a highly effective and safe imaging technique for evaluating vascular structures without using contrast agents. We propose in this work an automated analysis of the acquired NE-MRA to extract the thoracic aortic shape in 3D and the computation of its morphology. Quantitative description of morphology in the whole thoracic aortic shape supports a fast and precise prophylactic surgery decision as well as the evaluation of thoracic aortic determinants that impact the morphology. Beyond investigation of correlations between determinants and morphological changes in the thoracic aorta, the identification of causal relationships is essential for effective diagnoses and therapy planning. Since correlation does not imply causation, confounding factors may exist that create spurious correlations which lead to wrong conclusions and biased diagnosis. Hence, causal investigations are indispensable to identify the causal determinants towards morphological changes in the thoracic aorta.

## INTRODUCTION

The thoracic aorta, which is part of the aortic organ^1^, is a vital component of the circulatory system, serving as the main conduit for systemic blood perfusion originating from the heart. Understanding thoracic aortic pathology is essential, given its susceptibility to a spectrum of pathological changes.^1^ Such changes range from progressive aneurysmal dilation to life-threatening aortic dissection. In addition to genetically triggered causes such as Marfan syndrome and Ehlers-Danos syndrome, male sex, age, body surface area (BSA), hypertension, and alcohol and tobacco consumption have been reported as potential risk factors for thoracic aortic dilatation^2–7^, while diabetes is reported as a factor that decreases the risk for dilatation of the thoracic aorta.^8,9^ However, in the general population, distribution and determinants of the thoracic aorta are inadequately studied due to a lack of large population-based studies with adequate imaging of the aorta. The German National Cohort (NAKO), a large epidemiological, multicentric, prospective cohort study that comprises more than 200,000 randomly selected participants from the general German population, offers a unique opportunity to study aortic morphology and pathophysiology. In addition to epidemiological, functional, and laboratory data, the NAKO database includes MRI scans of 29,913 subjects acquired using a standardized imaging protocol. Non-contrast-enhanced magnetic resonance angiography (NE-MRA) acquisitions of the thorax are part of this protocol enabling dedicated examination of the thoracic aorta.^10^

The analysis of such large datasets requires automated processing of imaging data. Deep learning (DL) techniques have been successfully applied across various imaging modalities for automated segmentation of organs. Such techniques have been primarily used on computed tomography (CT) imaging of the thoracic aorta^11^ and only few studies describe fully automated shape analysis of the thoracic aorta based on MRI following EACTS/STS guidelines.^12,1^

Previous observational studies investigating the ascending aortic shape were often conducted on small datasets^3,5–7^ or were limited to subpopulations (e.g. higher age groups as UK Biobank study).^13–16^ Most of these studies primarily conducted conventional statistical analyses that examined correlations between individual risk factors and morphological changes in the thoracic aorta. Such analyses, however, fail to capture causal determinants of aortic morphology. Causal analyses can help uncover causal relations and thus provide informed hypotheses that can be further evaluated in interventional study settings. Conventionally, causal discovery is employed in longitudinal studies. However, recent advances have demonstrated the applicability of causal discovery methods to observational cross-sectional studies.^17–19^

The purpose of this study was to provide morphological characterization of the ascending aorta based on NE-MRA imaging volumes from the NAKO study and to investigate possible determinants of ascending aortic morphology. Our unique contributions include: (1) Fully automated, DL-based 3D shape analysis of the ascending aorta, (2) Descriptive statistical analyses of aortic phenotypes across the general population, and (3) Investigation of causal determinants of mid-ascending aortic diameter (AAoD) based on a large-scale cross-sectional study.

## METHODS

This study was approved by all study centers’ local ethics committees in accordance with the Declaration of Helsinki. Written informed consent was obtained from all participants. This work uses the notation mean ± standard deviation. All analyses were performed using Python (Python version 3.9.5)

### (1) Study population

The initial study population encompassed 29,913 participants with available MR imaging data acquired in NAKO study centers in Germany between 2014 and 2019. Of these, 329 datasets were excluded after image quality assurance, 4,503 datasets were excluded due to missing reported data entries, and 337 datasets were excluded due to unrealistic data entries (e.g. negative age, higher number of tobacco packyears than age), resulting in a total of 25,073 datasets used in this study (males: 13,851, females: 11,222, age: 47.7±12.3 y [19 y to 74 y]).

#### Data

Imaging data were acquired as non-contrast-enhanced T2-weighted free-breathing MR angiography of the thorax (details provided in Supplemental Methods, examples in Supplemental Figure 1). Each dataset included additional tabular information (acquisition details provided in Supplemental Table 1). Of these, we used the entries sex, age at examination, height, and weight, which were standardized collected or measured at baseline by trained personnel using a calibrated integrated measurement station (SECA model 764, Seca®, Hamburg, Germany), and computed the BSA using Dubois formula. Additionally, we included self-reported information on physician’s diagnosis of hypertension and diabetes obtained through an interview with the answering option “yes”/“no” as well as the self-reported frequency of the consumption of alcohol categorized as “never”, “<1/month”, “2-4/month”, “2-3/week”, “4-6/week” and “daily”. Tobacco consumption was quantified in pack-years (20 cigarettes per pack) computed from self-reported quantity and duration of tobacco consumption.

### (2) Shape analysis of the thoracic aorta

To obtain morphological shape measures of the thoracic aorta, NE-MRA were processed as depicted in Figure 1. In short, the thoracic aorta was segmented using a DL-based framework which additionally predicted the following six thoracic aortic landmark points: sinotubular junction (border between aortic root and ascending aorta), origin of brachiocephalic artery, left carotid artery, and left subclavian artery (with all three landmark points at the aortic wall in the center of each vessel’s branch), diaphragm (at the hiatus), and end of image. These outcomes were used to automatically determine the morphology of the aorta, including the length, volume, and diameter of the ascending aorta (Supplemental Material).

**Figure 1.**
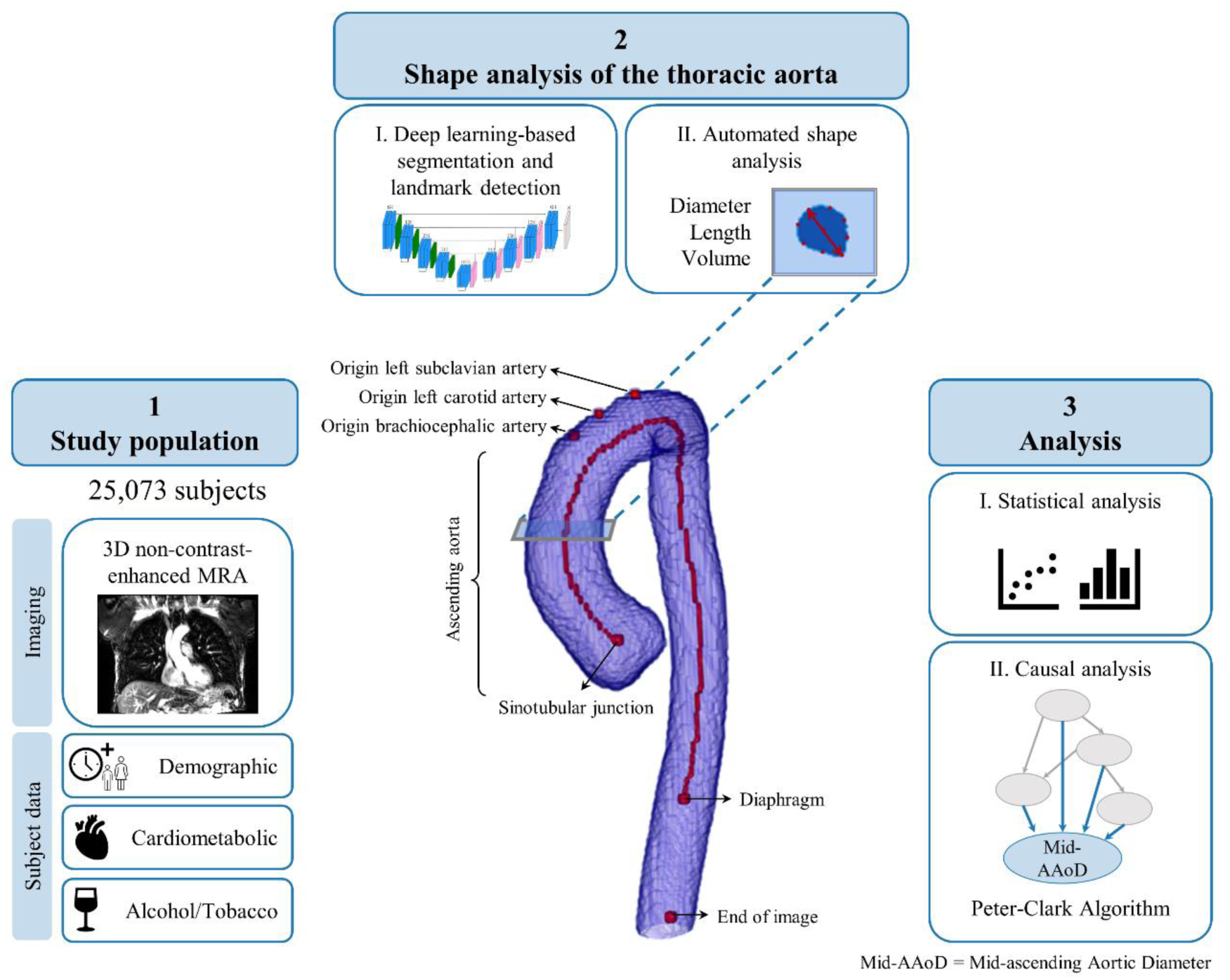
Workflow to analyze determinants of ascending aortic morphology. (1) Description of the NAKO study population including non-contrast-enhanced magnetic resonance angiography (NE-MRA) of 25,073 subjects. (2) Automated description of the thoracic aorta from NE-MRA. Using deep learning for the 3D segmentation and automatically computing based on the extracted segmentation mask the thoracic aortic (maximal) mid-AAoD, length, and volume (3) Standard statistical analysis of demographic (sex, age, height, BSA) and cardiometabolic (hypertension, diabetes) factors, as well as alcohol and tobacco consumption and the mid-AAoD. Proof-of-concept of an advanced causal analysis to go beyond statistical tests and give insights into the actual causal

#### I. Deep learning-based segmentation and landmark point detection

The DL model (Supplemental Figure 2) is an extension of a previously published segmentation model^20^ which is based on a residual UNet architecture. It automatically predicts the segmentation mask of the thoracic aorta and the above-described six landmark points (details in Supplemental Methods).

#### II. Automated shape analysis

The thoracic aortic shape was automatically analyzed by first computing the centerline, followed by the computation of the length and volume of the ascending aorta. The maximum aortic diameter was automatically extracted at 200 vessel cross-sections along and perpendicular to the centerline (details in Supplemental Methods). The following analysis used the mid-AAoD measured at the midpoint between sinotubular junction and origin of the brachiocephalic artery.

### (3) Statistical and causal analysis

The Python library SciPy (version 1.8.1) was employed for statistical analysis and causal-learn (version 0.1.2.8)^21^ for causal analysis.

#### I. Statistical analysis

Statistical tests, including univariate linear regression and stratified subgroup analyses adjusted for sex, were performed to assess relationships between mid-AAoD and the observed variables. All statistical tests were conducted using a significance level of 0.05. The coefficient of determination (r²) and the corresponding 95%-Confidence Interval (CI) were reported to quantify the strength of the relationships.^22^ Furthermore, multivariable Lasso regression was applied to investigate the impact of all observed variables on the mid-AAoD.

#### II. Causal analysis

The presence of spurious correlations due to confounding factors carries the risk of mistakenly assuming causal relationships. Conventional statistical tests can detect correlations but do not distinguish between direct causal dependencies and spurious correlations.

As part of this study, we provide proof-of-concept for causal discovery on observational cross-sectional data and investigate potential causal determinants of the mid-AAoD. For this investigation, we included reported variables sex, age, height, and BSA as well as information about prevalence of hypertension and diabetes, alcohol consumption, and pack-years of tobacco consumption history as possible causal determinants of the AAoD.

Causal dependence between the mid-AAoD and the respective variables was assessed using the Peter-Clark algorithm (PC)^18,23^. The output of PC is a causal graph that demonstrates the causal relationships between all observed variables with arrows pointing from the cause to the effect.

PC defines causal relationships based on conditional independence testing on observational cross-sectional data under the Causal Markov Condition. It assumes that two variables are causally dependent if and only if conditioning on any subset of remaining variables does not make them independent, following Reichenbach’s common cause principle^24^. The algorithm assumes Causal Sufficiency, indicating that there are no unobserved confounders. Additionally, the algorithm adheres to the Faithfulness Condition, which assumes that certain statistical relationships in a distribution imply corresponding graphical d-separations in the causal graph.

Dependencies between observed variables that were a-priori assumed to be causally unrelated, e.g., height not causally influencing sex, were precluded from consideration, while known causal dependencies, e.g., sex influences height, were inserted as required relationships. The background information for graph computation is listed in Supplemental Table 2.

## RESULTS

### (1) Study population

Among the 25,073 applied subjects of the NAKO database, 93.9% identified themselves as Caucasian, other ethnicities collectively represent ∼1%, and ∼5% did not specify their ethnicity. The 13,851 (55%) male subjects and 11,222 (45%) female subjects were distributed over an age range of 19 to 74 years. Females were on average slightly older than males (male: 47.2±12.4 y (95%-CI: 47.0-47.4), female: 48.3±12.2 y (95%-CI: 48.1-48.6)), and had on average a smaller body height and BSA (height: male: 179.2±7.0 cm (95%-CI: 179.1-179.3), female: 165.6±6.5 cm (95%-CI: 165.5-165.7), BSA: male: 2.05±0.17 m² (95%-CI: 2.04-2.05), female: 1.78±0.17 m² (95%-CI: 1.77-1.78)) (Figure 2A). Diagnosed hypertension was reported by 24.4% of male subjects and 21.1% of female subjects and diabetes by 3.9% and 4.0%, respectively (Figure 2B, 1C). Males tended to consume alcohol more frequently than females and showed higher lifetime tobacco exposure (7.0±12.6 pack-year (95%-CI: 6.8-7.2)) more frequently than females (4.8±10.0 pack-year (95%-CI: 4.6-5.0) (Figure 2D, 2E).

**Figure 2.**
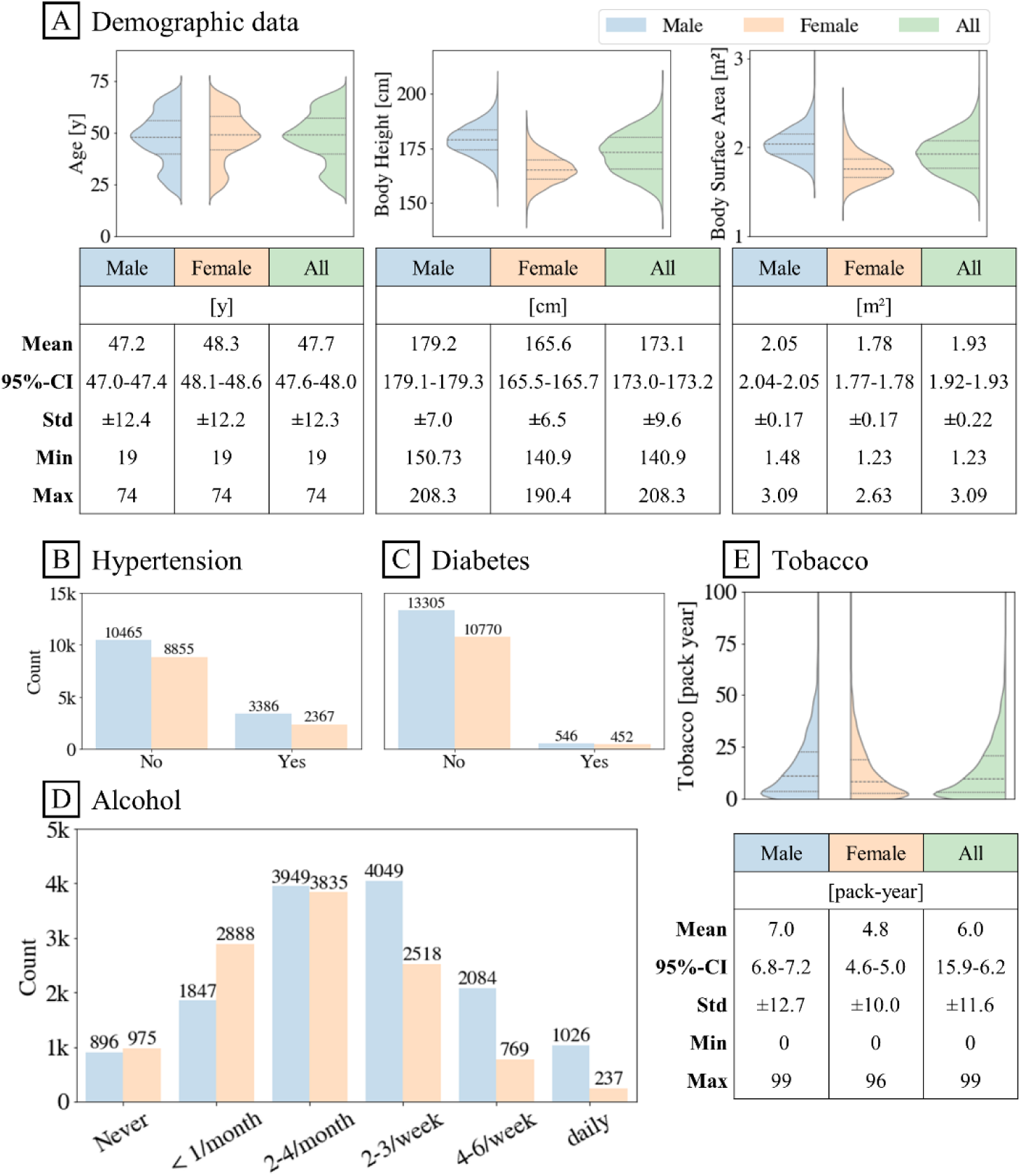
Epidemiological cohort characteristics of 25,073 subjects of the German National Cohort (NAKO). (A) Age (left), height (middle), body surface area (right) distribution of male, female, and all subjects. (B)/(C) Prevalence of hypertension/diabetes, (D) Alcohol consumption in frequency, (E) Tobacco consumption in pack-year by sex.

### (2) Shape analysis of the thoracic aorta

The mean length, volume, and mid-AAoD of the ascending aorta were larger in males than in females (Figure 3A). In males, the mean ascending aortic length was 8.1±0.8 cm (95%-CI: 8.1-8.1), the mean ascending aortic volume was 81.7±21.4 mm³ (95%-CI: 81.3-82.1), and the mid-AAoD was 35.6±5.1mm (95%-CI: 35.5-35.6). In females, the ascending aorta had a length of 7.4±0.8 cm (95%-CI: 7.3-7.4), a volume of 63.0±16.9 mm³ (95%-CI: 62.7-63.3), and a mean mid-AAoD of 33.3±4.5 mm (95%-CI: 33.3-33.4).

**Figure 3.**
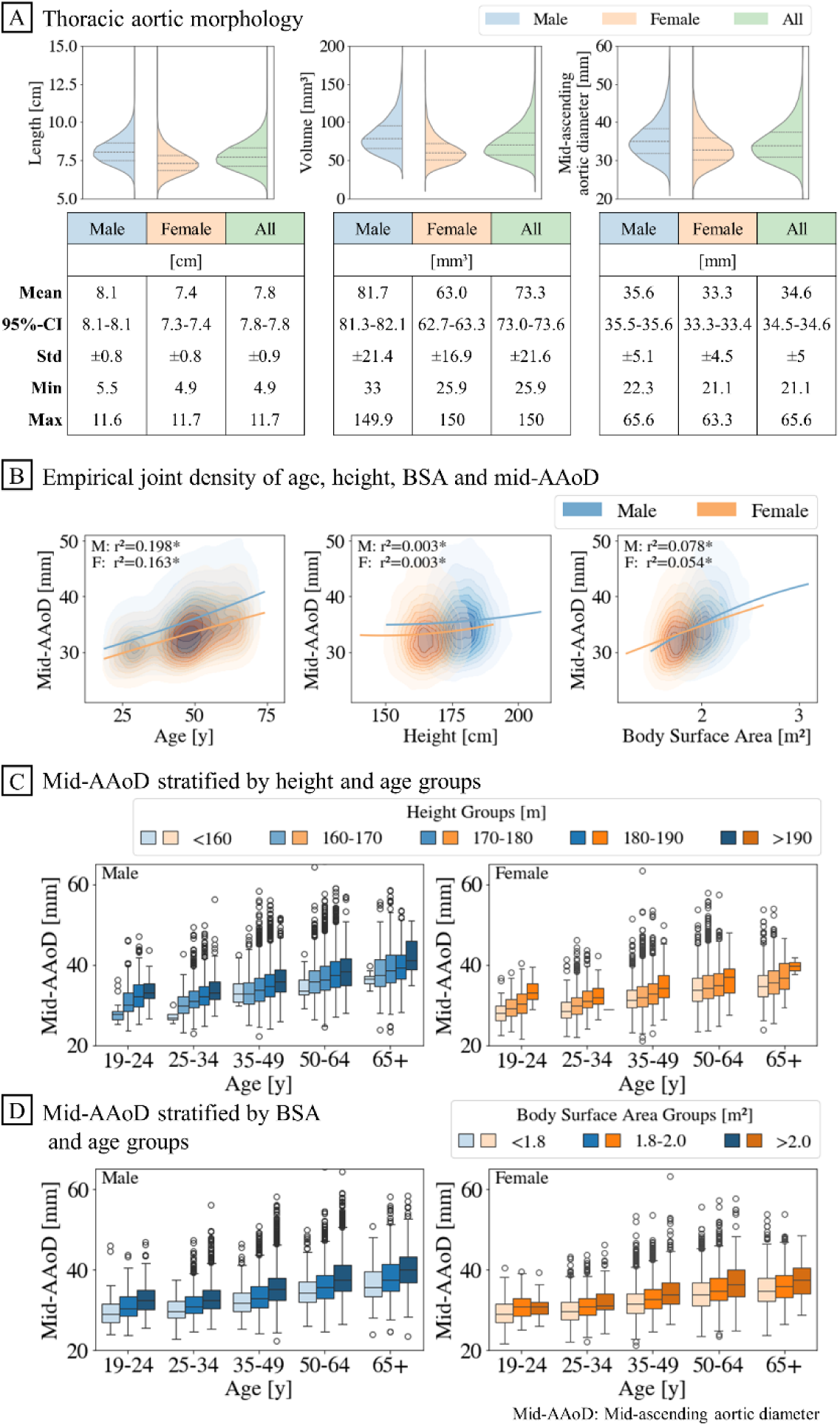
Distribution of the ascending thoracic aortic shape. (A) Automatically extracted length (left), volume (middle), and mid-AAoD (right) of the ascending aorta. Male and female values differed significantly. (B) (A) Age (left), height (middle), and body surface area (BSA) (right) and mid-AAoD by sex. Lines represent quadratic regression. (* Linear regression results, F: Female, M: Male) (C) Height, (D) BSA compared to the mid-AAoD within age groups by sex. Mid-AAoD increased in the presence of age, BSA, and height.

### (3) Statistical and causal analysis

#### I. Statistical analysis

##### Sex, age, height, and BSA

We found a moderate coefficient of determination between mid-AAoD and age (male: r²=0.198, F(1,13849)=3415.7, p<0.001, female: r²=0.163, F(1,11220)=2181.8, p <0.001), as well as with BSA (male: r²=0.078, F(1,13849)=1168.5, p<0.001, female: r² = 0.05, F(1,11220)=643.3, p<0.001) (Figure 3B). Between mid-AAoD and body height, the coefficient of determination was neglectable small (male: r²=0.003, F(1,13849)=41.8, p<0.001, female: r²=0.003, F(1,11220)=35.1, p<0.001). Besides the influences on the mid-AAoD, height decreased with age (male: r²=0.053, F(1,13849)=782.3, p<0.001, female: r²=0.07, F(1,11220)=861.2, p<0.001), while age and BSA showed no linear dependency (male: r²=0.0, F(1,13849)=1307, p=0.253, female: r²=0.0, F(1,11220)=1206, p=0.27) (Supplemental Figure 3).

###### Stratified analyses

Observing the mid-AAoD based on stratified groups with age divided into five groups: “19-24 y”, “25-34 y”, “35-49 y”, “50-64 y”, and “65+ y”, height into five groups: “<160 cm”, “160-170 cm”, “170-180 cm”, “180-190 cm”, and “>190 cm”, and BSA into three groups: “<1.8 m²”, “1.8-2.0 m²”,”>2.0 m²”, allowed to investigate the relationships within and across subgroups. Thereby, mid-AAoD increased independently with age, BSA, and height group (Figure 3C, 3D).

##### Hypertension and diabetes

Hypertensive and diabetic subjects showed larger mid-AAoD in both sexes compared to participants without the respective health condition (Figure 4A, 5A). Both, hypertensive and diabetic subjects, were on average older by 9.6 y (95%-CI: 9.5-9.7) and 8.8 y (95%-CI: 8.3-9.3) and had higher BSA by 0.082 m² (95%-CI: 0.080-0.085) and 0.07 m² (95%-CI: 0.058-0.081), respectively (Supplemental Table 3, 4).

**Figure 4.**
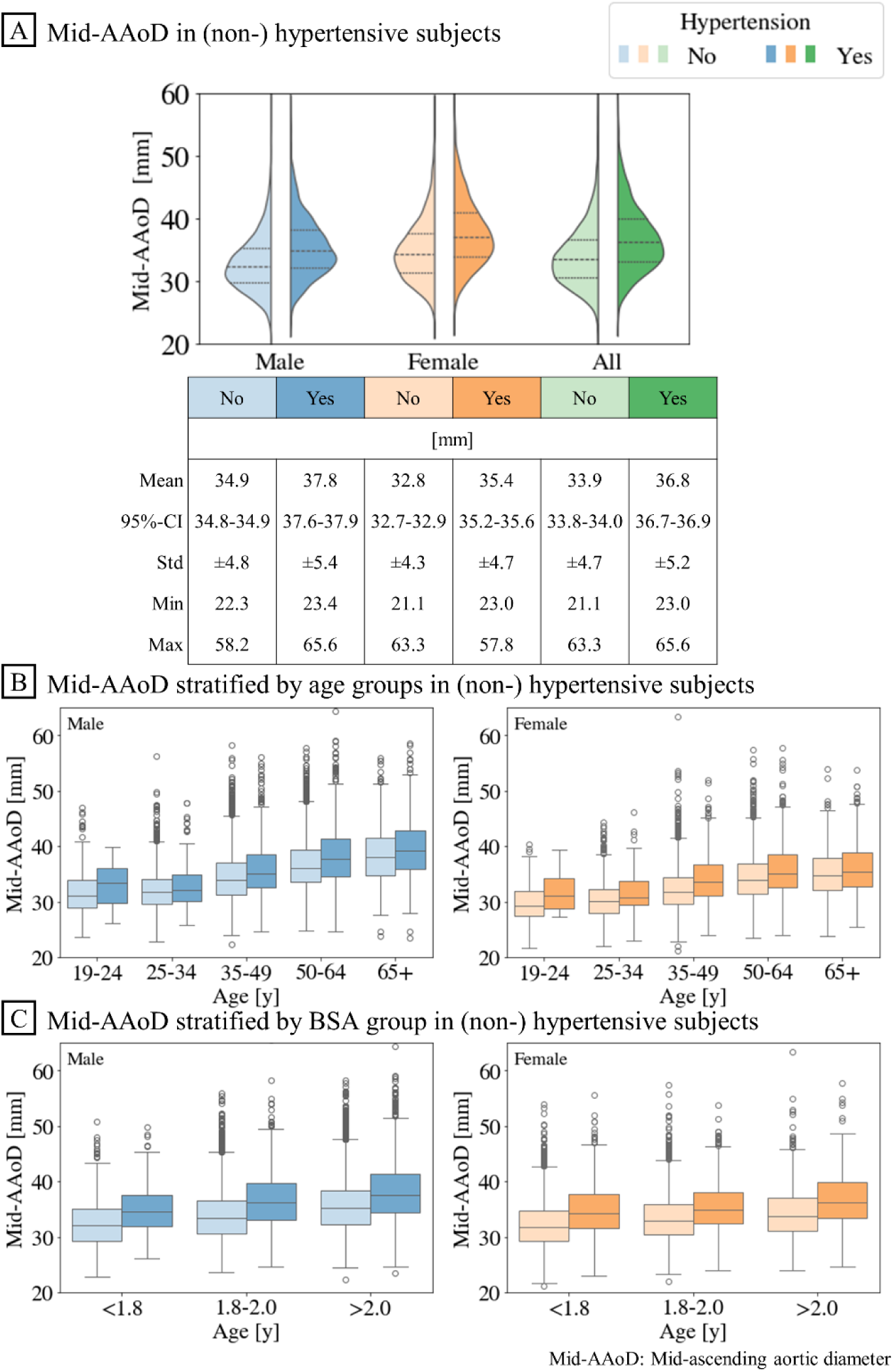
Relationship between hypertension and mid-AAoD. (A) Empirical distribution of the mid-AAoD in hypertensive and non-hypertensive subjects. (B)/(C) Diameter comparison of (non-)/hypertensive subjects stratified by age/body surface area (BSA) groups. Diameter increased with age and BSA as well as with the prevalence of hypertension.

**Figure 5.**
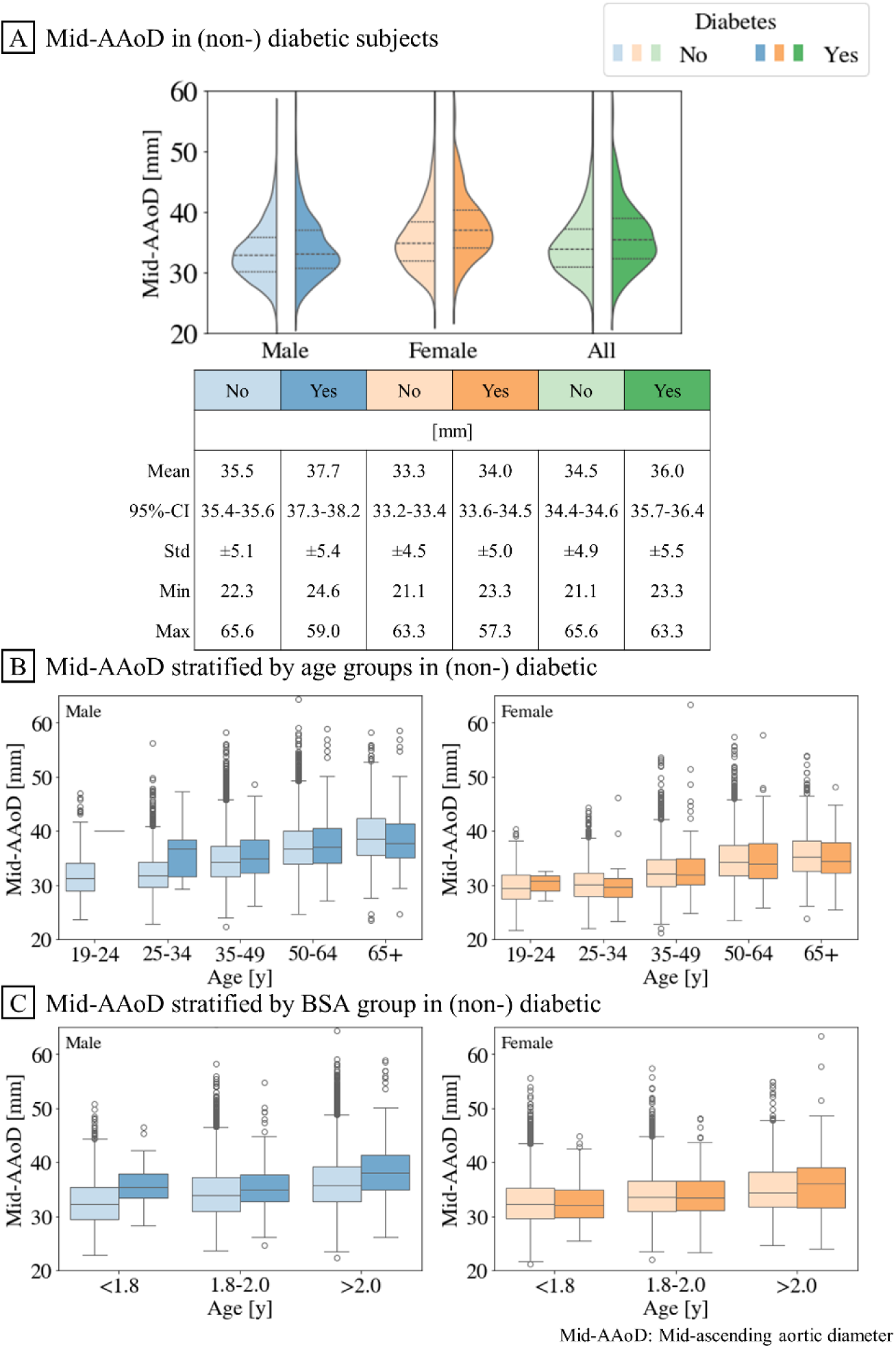
Relationship between diabetes and mid-AAoD. (A) Empirical distribution of the mid-AAoD in diabetic and non-diabetic subjects. (B)/(C) Diameter comparison of (non-)/diabetic subjects stratified by age/body surface area (BSA) groups. Diameter increased with age and BSA but not with the diagnosed diabetes.

###### Stratified analyses

In the age-stratified subgroup analysis, individuals with hypertension had larger mid-AAoD than non-hypertensive subjects across all age subgroups. No distinct differences in mid-AAoD were observed between diabetic and non-diabetic subjects across the age groups (Figure 4B, 5B). In the BSA-stratified analysis, among each BSA subgroup, both hypertensive and diabetic subjects showed larger mid-AAoD (Figure 4C, 5C).

##### Alcohol consumption

Subjects who reported “daily” alcohol consumption showed largest mean mid-AAoD in both sexes. We observed an absolute increase in mean mid-AAoD of 2.2 mm in males and 1.9 mm in females when comparing the consumption category “never” and “daily” (Figure 6A, Supplemental Table 5, 6).

**Figure 6.**
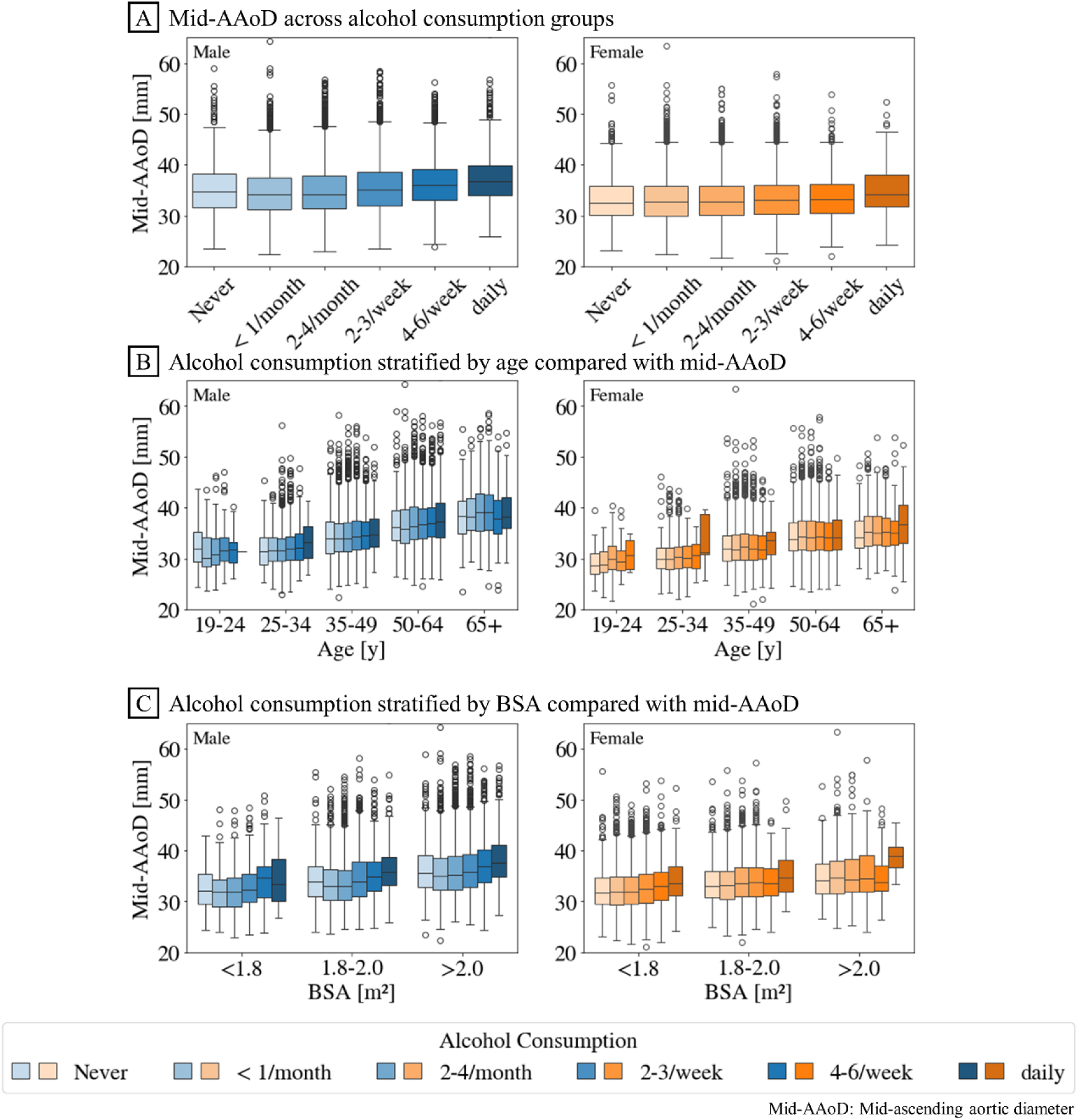
Relationship between alcohol consumption and mid-AAoD. (A) Distribution of mid-AAoD in each alcohol consumption category by sex. (B)/(C) Stratified distribution of mid-AAoD in each alcohol consumption category of different age/body surface area (BSA) groups. Mid-AAoD increased with age and BSA. In males, independent of age and BSA, mid-AAoD increased with an increased alcohol consumption.

In males, we observed a trend of increasing mean mid-AAoD with rising alcohol consumption. However, the 95%-CIs for the first three consumption groups overlapped. Significant differences were only observed in the higher consumption categories starting from “2-3/week”. In contrast, the increase in mean mid-AAoD in females was marginal, with an enlargement of 0.6 mm from the category “never” to “4-6/week”. Only the mid-AAoD of “daily” consumption showed a non-overlapping 95%-CI.

Generally, individuals with “daily” alcohol consumption also had the highest mean age with a difference between the categories “never” and “daily” of 8.6 y for males and 7.4 y for females. Additionally, we found that individuals who consumed alcohol more frequently also had a higher prevalence of hypertension. Among males and females who reported “daily” alcohol consumption, the prevalence of diagnosed hypertension was 36.5% and 30.0%, respectively, compared to 25.0% and 23.5% among males and females who reported “never” consuming alcohol (Supplemental Table 5, 6).

###### Stratified analyses

We observed that older subjects tended to have larger mid-AAoD regardless of their drinking habits (Figure 6B). Moreover, subjects with a BSA>1.8 m² and “daily” alcohol consumption exhibited higher mean mid-AAoD compared to those with lower alcohol consumption (Figure 6C).

##### Tobacco consumption

Higher lifetime tobacco exposure was associated with a slight increase in mid-AAoD in males (male: r²=0.012, F(1,13849)=162.35, p<0.001) and almost no increase in females (female: r²=0.002, F(1,11220)=24.76, p<0.001) (Figure 7A).

**Figure 7.**
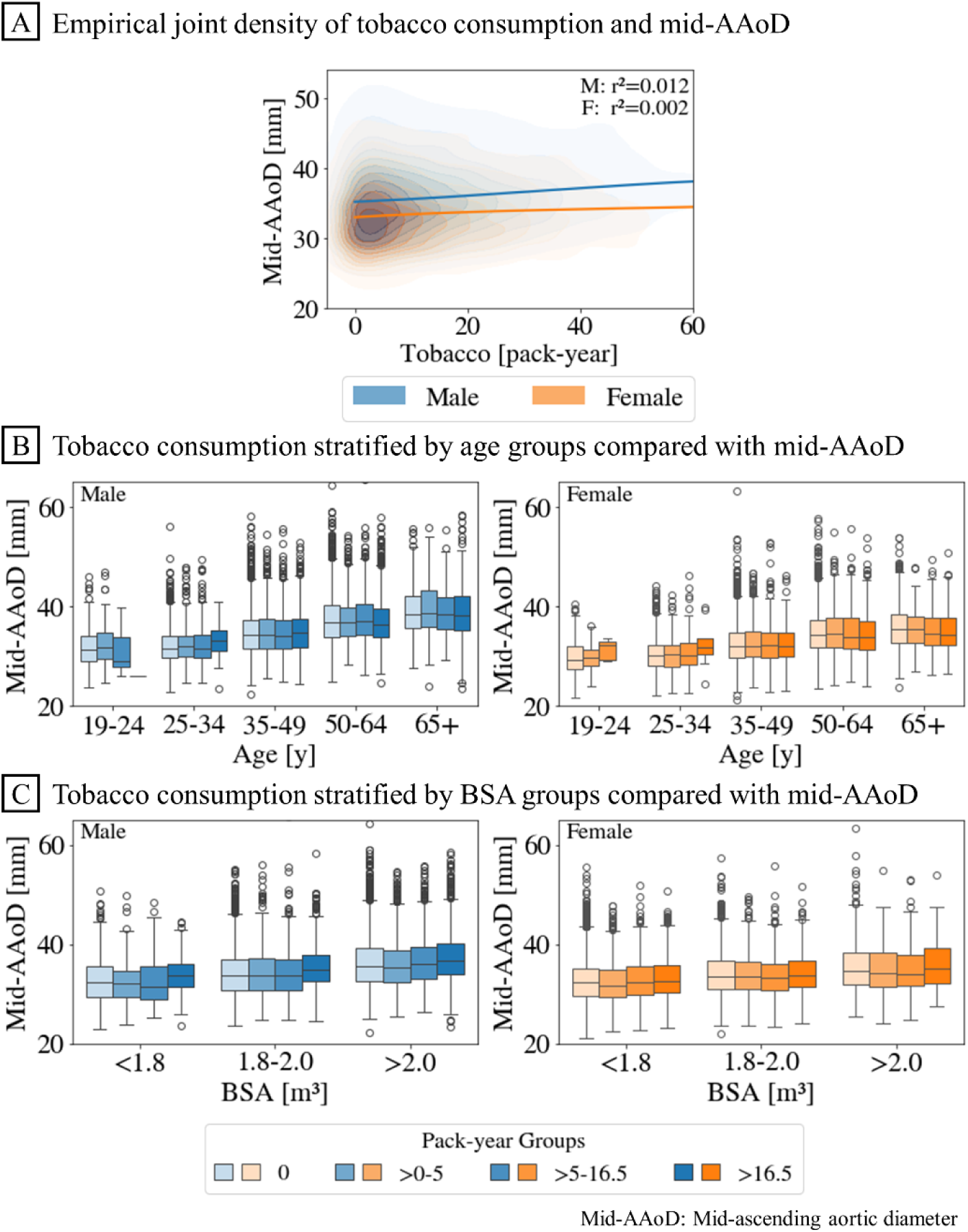
Relationship between tobacco consumption and mid-ascending diameter. (A) Empirical joint densities of mid-AAoD and tobacco consumption in pack-years by sex excluding subjects that never smoked. (B)/(C) Diameters in each tobacco group of stratified by age/body surface area (BSA) groups. The mid-AAoD was largest in highest tobacco consumption “>16.5 pack-year” when observed independent of BSA. No clear trend was identified in the influence of tobacco consumption when stratified by age.

By categorizing tobacco consumption into the pack-year subgroups “0”, “>0-5”, “>5-16.5”, and “>16.5”, we observed largest mid-AAoD in males and females reporting highest lifetime tobacco exposure of “>16.5 pack-years”, with 36.6±5.0 mm (95%-CI: 36.4-36.8) and 33.9±4.3 mm (95%-CI: 33.6-34.1), respectively (Supplemental Table 7, 8). The mean mid-AAoD of other pack-year groups showed overlapping 95%-CIs, indicating no distinct differences.

Subjects of the category “>16.5 pack-years” were the oldest with a difference of 6.0 y in males and 5.1 y in females compared to the next lower pack-year group, “>5-16.5 pack-years”, which was the second oldest group (Figure 7B). Additionally, within the “>16.5 pack-years” category, the prevalence of hypertension was also highest, showing an absolute percentage increase of more than 9% and 6% compared to the other categories in males and females, respectively. Hence, although individuals within the category “>16.5 pack-years” exhibited the highest mid-AAoD, this group also tended to be older on average, had a higher average BSA, and a higher prevalence of hypertension.

###### Stratified analyses

Stratifying the mid-AAoD by BSA demonstrated an increasing trend towards the highest lifetime tobacco exposure category “>16.5 pack-years” (Figure 7C). However, observing mid-AAoD stratified by age, revealed that in females, particularly in the higher age groups, mid-AAoD was smaller with higher lifetime tobacco exposure compared to the lower exposure categories. In males, no distinct trend was identified.

##### Multivariable analysis with all observed variables

The multivariable analysis with Lasso including all observed variables (Supplemental Figure 4) showed a statistical association of age, height, BSA, and hypertension on mid-AAoD, with age having the strongest effect with a coefficient of 1.94, followed by BSA with an impact of 1.5, hypertension with 0.24 and a small effect of height with 0.13. Lasso regression indicated no statistical effects of sex, diabetes, alcohol, and tobacco consumption on the mid-AAoD.

#### II. Causal analysis

Above-described stratified subgroup analyses demonstrated potentially spurious interactions between observed variables. While conventionally causal discovery is performed on longitudinal studies, PC algorithm allowed causal discovery in observational cross-sectional studies. The resulting causal graph by PC algorithm revealed causal determinants on the mid-AAoD as well as of the entire system of causal relationships between all observed variables (Figure 8).

**Figure 8.**
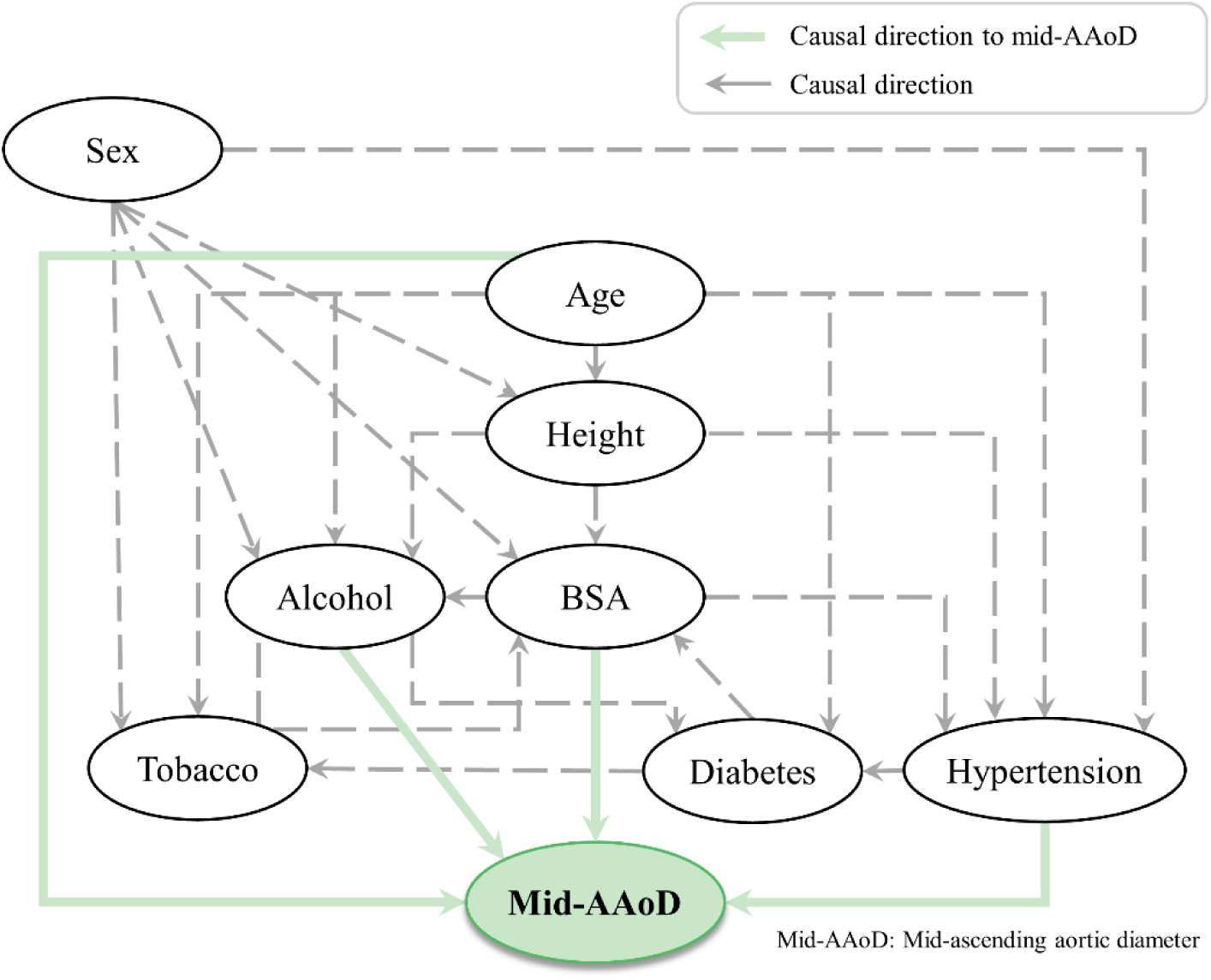
Causal graph between all observed variables and the mid-ascending diameter. Causal graph with all observed variables; Age and BSA showed causal influence on the mid-AAoD. Regarding the cardiometabolic factors: Diabetes was independent of mid-AAoD, whereas hypertension showed an influence on mid-AAoD. Alcohol showed impact on diameter, while tobacco was independent of mid-AAoD.

##### Sex, age, height, and BSA

Causal discovery indicated no causal relationship between sex and the mid-AAoD. As hypothesized, it revealed causal relationships between age and BSA and the mid-AAoD. The presumed relationship between height and mid-AAoD was not detected. Obtained causal graph also gave insights into the dependencies between age, height, and BSA, showing that age has a causal impact on height as well as height on BSA.

##### Hypertension and diabetes

Causal analysis suggested a direct impact of hypertension on the mid-AAoD. No direct causal relationship between prevalence of diabetes and mid-AAoD was observed. Besides the impacts on the mid-AAoD, hypertension appeared to causally influence diabetes.

##### Alcohol and tobacco consumption

A causal impact of alcohol consumption on mid-AAoD was observed, while no causal relationship was found between tobacco consumption and mid-AAoD. A bidirectional causal dependency (relationship without direction) was observed between alcohol and tobacco consumption.

## DISCUSSION

### (1) Study population

In this study, we investigated ascending aortic phenotypes in a large epidemiological imaging study (NAKO), which included NE-MRA of 25,073 subjects from the German population aged 19 to 74 y, addressing an important research gap identified by EACTS/STS^1^.

### (2) Shape analysis of the thoracic aorta

Previous studies performed automated analysis of the thoracic aorta either on CT data^25,26,11,27^ or only analyzed single points instead of describing the whole thoracic aorta^14,15,13^. To overcome these limitations, we improved our previous DL-based framework^20^ for three-dimensional segmentation of the thoracic aorta in NE-MRA by simultaneously predicting landmark points on the thoracic aorta. Hence, we were able to partition the thoracic aorta and to define the level of the mid-AAoD precisely according to EACTS/STS guidelines.^12,1^

We observed mean mid-AAoD of 35.6±5.1 mm and 33.3±4.5 mm in males and females, respectively, which is slightly larger than in previous large-scale European studies.^14,15,28^ However, in our study, we precisely extracted the maximal mid-AAoD, considering that the dilatation of the thoracic aorta can take on any irregular shape rather than a perfect geometric shape (circle, ellipse). Pirrucello^14,15^ reported mean AAoD of 33.2±3.4 mm (male) and 30.4±3.1 mm (female) in 40,363 cardiac MRI scans (UK Biobank), limited to ages 40 to 69 y. Their measurements were taken at the level of the right pulmonary artery, assuming that the aortic shape is an ellipse where the minor axis at its maximum size of a cardiac cycle determines the AAoD. Another MRI-based study of 3,573 multiethnic subjects found mean AAoD of 32±4 mm, with no subject exceeding a AAoD of 50 mm, providing only a restricted perspective on the thoracic aortic morphology.^29^ In contrast, Asian population-based studies showed smaller AAoD on cardiac ultrasound and CT, reporting mean sizes of 28.1±3.2 mm (647,087 subjects)^30^ and 29.9±5.7 mm (300 subjects)^31^, respectively. Non-contrast-enhanced CT in 2,952 American subjects (mean age: 55.0±10.2 y) with AAoD measurements at lower level of the pulmonary artery bifurcation yielded 34.0±4.1 mm in males and 31.8±3.7 mm in females.^32^ Similar results were observed in CT angiographies from the Copenhagen General Population study (902 subjects, age: 40-80 y) with AAoD of 33±4 mm in males and 30±3.5 mm in females. However, the study excluded subjects with cardiovascular risks, including diabetes and hypertension, where the latter showed an impact on the AAoD in our study.^28^

In this study, we derived the maximum mid-AAoD according to the EACTS/STS guideline^1^, enabling precise measurements regarding position and technique. By encompassing a broad age range and incorporating all subjects without restrictions, our results uncovered mid-AAoD of the general population. The provided framework allows to extract aortic diameters on 200 points along the aorta, which can support prophylactic surgery decision based on the absolute largest ascending aortic diameter.

### (3) Statistical and causal analysis: Determinants of ascending aortic morphology

Beyond well-established associations between sex, age, BSA, and mid-AAoD^33,34,3^, we observed positive statistical associations between the mid-AAoD, self-reported hypertension, self-reported diabetes, and alcohol consumption. Simple correlation analysis revealed no substantial correlation between mid-AAoD and body height. Moreover, the statistical findings did not allow the drawing of any definitive conclusions regarding the tobacco influence on the mid-AAoD.

Stratified subgroup analyses and causal discovery analyses as well as multivariable Lasso regression revealed that these observed associations were in part confounded reflecting spurious correlations.

Although males showed significantly larger mid-AAoD than females, no causal relationship between sex and mid-AAoD was found, suggesting that the difference is likely confounded by height and BSA. As shown in other studies^35,13,36^, we observed a larger mid-AAoD in hypertensive subjects independent of age and BSA. Causal analysis confirmed direct causal relations between hypertension and mid-AAoD. However, causal analysis revealed no direct causal effect of self-reported diabetes on the mid-AAoD – the observed correlation was confounded by age and BSA. Consequently, causal analysis did not substantiate the previously suggested protective influence of diabetes regarding ascending aortic dilatation.^9,37,38^

A causal effect of alcohol consumption on mid-AAoD was confirmed. The multivariable Lasso analysis neglected the influence of alcohol, most likely due to the stronger impact of age, BSA, and hypertension. Age and hypertension, both clearly confounded the observed mid-AAoD of the single alcohol consumption categories, especially observable for the category of “daily” consumption, where the subjects were significantly older and had larger BSA. The observed increasing mid-AAoD trend with alcohol consumption was stronger in males than in females. It is unclear whether this difference is due to differences in biological responses to alcohol consumption, due to uncontrolled effects such as differences in self-reporting of alcohol consumption or amount of alcohol consumption.

Previous studies have been inconclusive on the tobacco consumption’s effect on the AAoD.^2,39^ PC algorithm and multivariable Lasso regression did not identify a direct causal effect of tobacco consumption with pack-years on mid-AAoD. This confirms that the apparent weak correlation was confounded by BSA, given that pack-years were correlated and showed a causal relationship with BSA. In addition, our causal analysis revealed a bidirectional relationship between alcohol and tobacco. Both factors are known as comorbidity,^40^ and a definitive causal direction remains indeterminate.

#### Study limitations

The present study assessed relationships of mid-AAoD and selected reported variables. However, additional factors may impact the mid-AAoD. Excluding factors might result in spurious correlations, creating relationships between variables that would otherwise be independent in the presence of a confounder. While it is possible to include additional factors in the causal analysis, this would lead to more complex graphs, as the causal graph provides insight into the entire system of causal dependencies between all observed variables. The PC algorithm does not quantify the strength of relationships, thus providing no confidential measure to assess uncertainties.

The variables hypertension, diabetes, alcohol, and tobacco consumption are self-reported by the subjects and not revised by experts. This introduces noise into the data, potentially leading to inaccurate representations of the actual current state. Additional data on blood pressure and vascular stiffness could not be included due to singular measurements, limiting their ability to represent ongoing changes.

The results of this study are based on observational cross-sectional data at one time point and are not validated within a clinical study. Imaging was only acquired once, hence precluding observation of longitudinal changes over time.

#### Outlook

In the scope of the NAKO study, follow-up whole-body MRIs are currently being acquired to provide a second examination point of subjects which will enable longitudinal causal evaluation. Furthermore, future research comprises broadening the DL analysis and causal proof-of-concept to the entire thoracic and abdominal aorta.

## CONCLUSIONS

Ascending aortic morphology varies significantly within the large epidemiological NAKO population depending on demographic factors, individual health status, and consumption of alcohol and tobacco. Automated DL analysis with full and precise description of aortic morphology is possible using NC-MRA acquisitions. Causal analysis of cross-sectional data can assist in disentangling observed correlations, identifying potential causal determinants of ascending aortic morphology, and providing insights into the overall system of causal dependencies within a study. Our causal analysis revealed that sex, age, BSA, hypertension, and alcohol consumption are causally related to the mid-ascending aortic diameter.

## Data Availability

The data that support the findings of this study are available from the German National Cohort but restrictions apply to the availability of these data, which were used under license for the current study, and so are not publicly available.

## ABBREVIATIONS

AAoD: Ascending Aortic Diameter
BSA: Body Surface Area
CI: Confidence Interval
CT: Computed Tomography
DL: Deep Learning
EACTS/STS: European Association for Cardio-Thoracic Surgery and The Society of Thoracic Surgeon
MRI: Magnetic Resonance Imaging
NAKO: German National Cohort
NE-MRA: Non-contrast-enhanced Magnetic Resonance Angiography
PC: Peter-Clark algorithm

## Acknowledgements

This project was conducted with data from the German National Cohort (NAKO) (www.nako.de). The NAKO is funded by the Federal Ministry of Education and Research (BMBF) [project funding reference numbers: 01ER1301A/B/C, 01ER1511D and 01ER1801A/B/C/D], federal states of Germany and the Helmholtz Association, the participating universities, and the institutes of the Leibniz Association. We thank all participants who took part in the NAKO study and the staff of this research initiative. We wish to disclose the use of the artificial intelligence tool DeepL for minor translations.

